# Accuracy of fingertip pulse oximeters: a device accuracy study

**DOI:** 10.64898/2025.12.05.25341372

**Authors:** Susannah Fleming, Matthew Titterington, Helen F Ashdown, Najib M Rahman

## Abstract

**Aim:** To assess the accuracy of commercially available pulse oximeters at various price points, and to investigate how long is needed to obtain a reliable measurement.

**Methods:** All 75 participants had pulse oximetry with 6 test oximeters and a reference (gold standard) oximeter. Where blood gases were clinically indicated, these results were also recorded.

**Results:** Compared to the reference oximeter, average errors in oxygen saturation ranged from 0.6% to 2.6% overestimation. All devices appeared to require some time to “settle” to an accurate reading, but accuracy at 30 seconds from application was similar to that at 2 minutes. There was no clear association between device cost and accuracy.

**Conclusions:** Fingertip pulse oximeters tended to overestimate oxygen saturation by up to 2.5% on average. We recommend that pulse oximeters are not read immediately after application, but are allowed at least 30 seconds to settle to an accurate reading.

**KEY MESSAGES:**

- Fingertip pulse oximeters are widely used in clinical care, particularly in the primary care and pre-hospital environment, and increasingly for self-monitoring by patients.
- We found that pulse oximeter accuracy varies for both oxygen saturation and heart rate, with 2.5% overestimation of oxygen saturation on average.
- Price is not necessarily an indicator of accuracy in pulse oximeters.
- Accuracy can be improved by waiting at least 30 seconds before recording oxygen saturation.

**PLAIN LANGUAGE SUMMARY:** Pulse oximeters are non-invasive fingertip medical devices that measure the amount of oxygen in the blood (“oxygen saturations” or “sats”), which in healthy people should be between 96% and 100%. They also measure heart rate. Some pulse oximeters are available to buy by the public quite cheaply, and their use for home monitoring has become more common during and since the COVID-19 pandemic. We wanted to understand if cheap oximeters are still accurate. We bought 6 different pulse oximeters and tested them on 26 healthy people and 49 patients who were attending a hospital outpatient clinic. We compared the oxygen saturation measurements from the 6 cheap oximeters with measurements from a hospital quality pulse oximeter as a “gold standard”. We compared the heart rate measurements from the pulse oximeters to a measurement of heart rate made by hand by a nurse. We also measured how long it took for the pulse oximeter to give an accurate measurement.

We found that the cheap pulse oximeters generally gave an oxygen saturation value that was between 0.6% and 2.6% higher than the hospital pulse oximeter. However, sometimes there was a larger difference of up to 7.4%. The cheap pulse oximeters also generally gave a heart rate that was between 0.8 and 2.7 beats per minute higher than the rate measured by a nurse. After 30 seconds, the pulse oximeter measurements were about as accurate as they were after 2 minutes. We found that very cheap pulse oximeters were just as accurate as less cheap pulse oximeters.

Our results suggest that cheap pulse oximeters tend to overestimate oxygen saturations and heart rate, but usually by a small amount which would not make much difference to the decisions a doctor would make. However, sometimes there may be a bigger difference so we would recommend repeating with a different device if there was concern about the patient.

## INTRODUCTION

Oxygen saturation (SpO_2_), measured using pulse oximetry, is now a standard vital sign, which is measured routinely in both hospital and community care, and forms part of many early warning scores, including the NEWS2 score.(1) Pulse oximetry became particularly important during the COVID-19 pandemic, when patients presented with low oxygen saturation but no apparent respiratory distress, and could be monitored remotely in clinician-led programmes and purchased by the public for self-monitoring. (2–7)

Assessments of the accuracy of pulse oximeters have typically focussed on the more expensive, validated devices used in hospitals.(8–13) However, oximeters used in primary and community care, and available for purchase by the public, are typically smaller and cheaper “fingertip” devices. It is not currently clear whether such devices are accurate, with varying results from existing studies.(14–16)

Despite the widespread use of pulse oximetry, there is little guidance on obtaining an accurate measurement. Observation by the authors and discussion with other clinicians suggested there is a “settling time” phenomenon, where the first measurement reported by the oximeter is not necessarily correct, and stabilises after a period of time. However, there is no evidence on this phenomenon, nor any guidance on how long to wait before a measurement could be considered “stable”.

We aimed to assess the accuracy of commercially available pulse oximeters at a range of price points, across a range of oxygen saturation levels, as well as to investigate the “settling time” phenomenon.

## METHODS

The full study protocol is provided as supplementary information to this paper.

### Test devices

We purchased six models of fingertip pulse oximeters (D1 to D6, see Supplementary Figure S1 and Supplementary Table S1, £22.95 to £284.96) from medical product suppliers. Devices were chosen to represent a range of price points and manufacturers, and be representative of the type of device in use in primary and community care, informed by results of a previous survey of UK General Practitioners (unpublished).

As a reference standard for oxygen saturation in all participants, we used the pulse oximetry unit Ohmeda 3700, with its standard connecting fingertip probe. This machine is widely used in UK hospitals, including in our specialist respiratory department where it is the standard-of-care, and it has previously been validated in a clinical research setting against arterial blood gas measurements.(17) Prior to this study it was tested by the hospital Clinical Engineering Department, and undergoes regular quality assurance checks as standard. For the subgroup of patients who underwent arterial or capillary blood gas analysis as part of their routine clinical care, we carried out a secondary analysis using this as the reference. As a reference standard for heart rate in all participants, we used manually recorded radial pulse carried out by a research nurse.

### Study population

This was a prospective study; we recruited both healthy volunteers and patients being assessed at the home oxygen clinic at Oxford University Hospitals (OUH) NHS Foundation Trust. The home oxygen clinic assesses patients for suitability for home oxygen therapy, as well as reviewing existing home oxygen users. Home oxygen therapy is when supplemental oxygen is provided in the patient’s home, due to hypoxaemia associated with chronic respiratory disease, so basing our study here meant we could recruit a cohort of participants with a range of blood oxygen saturations. Patients were provided with information about the study with their clinic invitation letter. Healthy volunteers were recruited from OUH and University of Oxford staff.

Participants were recruited consecutively between February and May 2015. Inclusion and exclusion criteria are provided in Supplementary Table S2. We aimed to recruit 25 “healthy” volunteers (defined as adults in good general health) and 50 participants from the home oxygen clinic (normal and lower SpO_2_ levels).

### Study procedure

For each participant, 3 test oximeters and the reference oximeter were applied to the four fingers of the right hand for two minutes (Figure 1). To allow for measurement of any “settling time” effect, all oximetry probes were attached to the patient’s fingers, and then the oximeters were turned on in as short a time as possible. Where a fingertip oximeter had no power button, it was attached at the same time as other oximeters were turned on. The process was repeated using the remaining 3 test oximeters and the reference oximeter, meaning each participant was tested with all 6 test oximeters and the reference oximeter was always used, resulting in 6 paired measurements per participant. Any supplemental oxygen use was recorded. A research nurse measured the radial pulse at the opposite wrist for 60 seconds during each two-minute period, for all participants.

**Figure 1.**
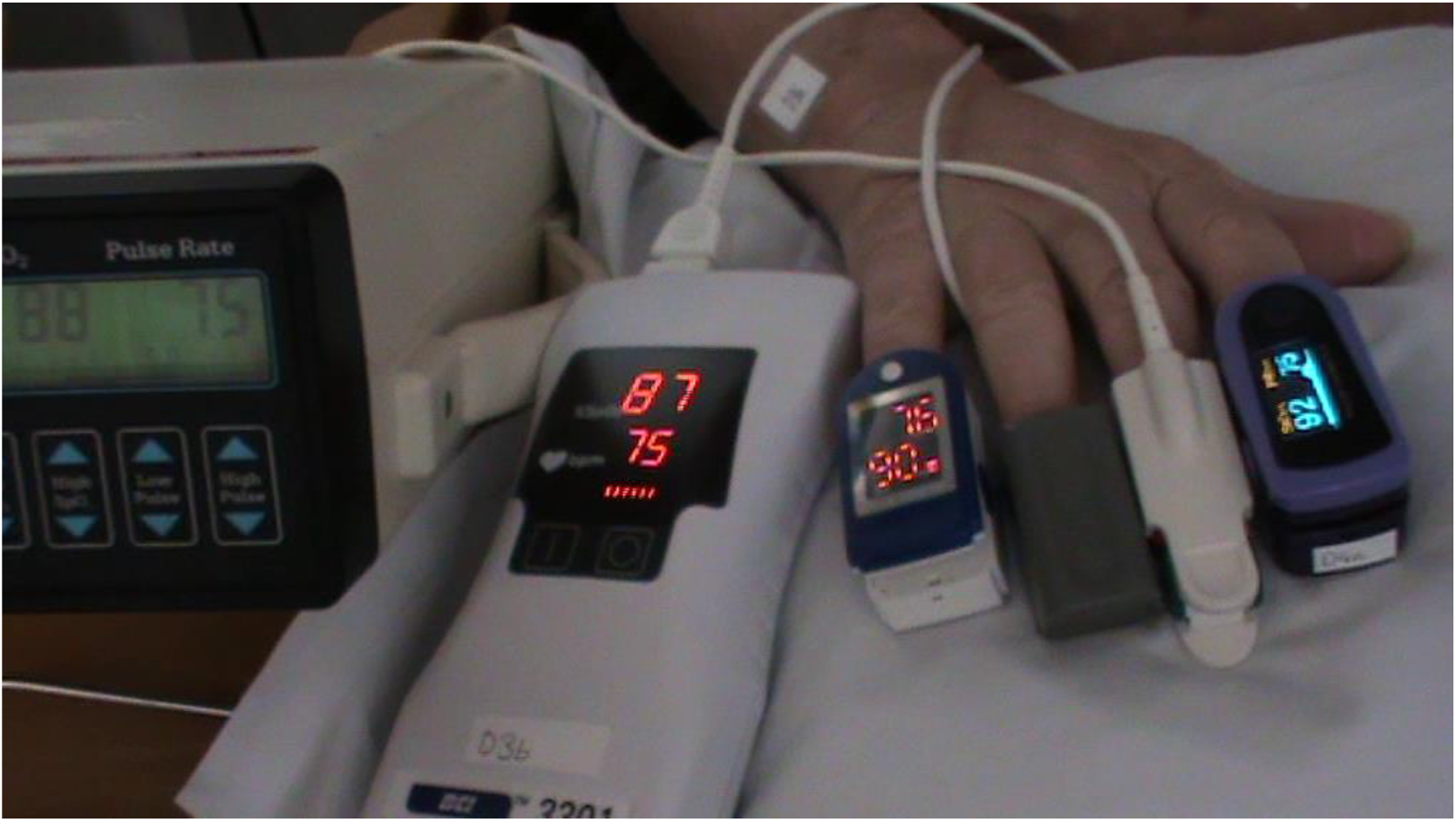
Still image from video recording of a data collection session, showing reference oximeter (far left), and three test oximeters (left to right: D3, D6 and D4). Patient consent to publish obtained.

If clinic participants required either capillary or arterial blood gas measurement as part of their standard care, we obtained permission to record oxygen saturations measurement from their sample. This was used for a sensitivity analysis using this as a reference standard instead of the reference oximeter. Oximetry measurements were carried out close to blood gas measurement, using the same levels of supplemental oxygen. Choice of capillary or arterial blood gas measurement was made by the treating clinician, and we were unable to obtain this information retrospectively.

### Randomisation

Each test oximeter was assigned a number (D1 through D6). For each patient, we randomised the order in which oximeters would be placed on the index to little fingers across the two measurement periods. To ensure no effect of individual device variation, two separate devices for each test oximeter were used, and we randomised which device would be chosen each time an oximeter was used.

Randomisation sequences were generated using random numbers obtained from random.org, carried out prior to any recruitment. Randomisation details were placed in sealed opaque envelopes labelled with sequential study numbers. Once a participant was recruited, the next envelope was opened.

### Data collection

We video-recorded the screens of the four pulse oximeters during each two-minute measurement period, in a single shot as shown in Figure 1. Data were extracted from the videos at a later date by two independent viewers, who were blind to the other viewer’s results, and to any blood gas results.

The independent viewers extracted the following data from the video recordings:

- Time of at which each oximeter was applied (or turned on, in the case of oximeters with a power button)
- Time of first reading and values displayed for both SpO_2_ and heart rate on each oximeter
- Time of first “good quality” reading on oximeter, along with SpO_2_ and heart rate displayed
- SpO_2_ and heart rate displayed on each oximeter at 15s, 30s, 45s, 60s, and 120s from initialisation of the last oximeter
- Participant skin colour (coded subjectively by the video viewer as light, intermediate, or dark).

“Good quality” readings were determined by either a clear pulsatile rhythm if the oximeter displayed a photoplethysmograph trace or a pulse bar, or by the change from red to green of the signal quality light, where present. If neither of these indications were provided by the device, it was assumed that a good quality reading was when both heart rate and SpO_2_ were first displayed together. We did not collect data on participant demographics other than subjectively assessed skin colour.

### Data quality

Data was accepted for analysis if the two independent viewers met a minimum acceptable level of agreement. These were defined as below based on clinical judgement and observation of variability in readings over short periods of time:

- Within 2% for SpO_2_ values
- Within 5bpm for heart rate values
- Within 6s for time to first good quality reading
- Within 3s for all other times

Only data where both viewers agreed within the above limits were retained in analysis. Where viewers agreed within the above limits, but did not completely agree, the average was used for analysis.

### Sample size

A sample size calculation was carried out based on a desired accuracy of ±1% for the 95% confidence intervals around the limits of agreement on the Bland-Altman plots.(18) We estimated the standard deviation of the agreement to be 2.5%, based on estimates from the literature of 2.1% and 3.1%, (8,19) and the usual quoted accuracy in the device leaflets of ±1-2%. This gave us a minimum sample size of 75 participants.

### Statistical analysis

Data were analysed using Bland-Altman analysis, with 95% confidence intervals calculated for both bias and 95% limits of agreement. For the primary analysis, differences were calculated between SpO_2_ results from oximetry, and the results for the reference oximeter (denoted as R). A secondary analysis compared SpO_2_ results from oximetry, and oxygen saturations from blood gas analysis, for the subset of patients where this was measured.

In order to investigate the “settling time” effect, we calculated the error at each time point from the value displayed on that oximeter at 120 seconds. These errors were also subject to Bland-Altman analysis for each time point on each device.

All analysis was carried out in R, with the *blandr* library being used to carry out Bland-Altman analysis.

## RESULTS

### Recruited patients

We recruited 75 participants: 49 from the home oxygen clinic, and 26 healthy volunteers. Median SpO_2_ from the reference device was 93% (IQR 90%-96%). Data from blood gas analysis was available for 45 home oxygen clinic participants. Sixteen patients were recorded as receiving supplemental oxygen during measurement. Subjective assessment of participant skin colour was assessed as “light” for 49 participants, “intermediate” for 7 participants, and “dark” for 2 participants. Subjective assessment of skin colour was inconsistent between video extractors for the remaining 17 participants. No adverse events were observed during the study.

### Data quality

For all SpO_2_ measurements extracted from video recordings, 90% of double-extracted data met our quality criteria for analysis. Complete agreement between viewers was found in 83.9% of SpO_2_ measurements, and 6.2% required averaging. Agreement was lower for heart rate measurements at 88.6%, with 66.7% of all double-extracted heart rate measurements in complete agreement, and 21.9% requiring averaging (Supplementary Table S3).

### Primary analysis

The primary analysis at 2 minutes showed that test oximeters typically overestimated SpO_2_ compared to the reference oximeter, with biases (average errors) ranging from 0.6% to 2.6% overestimation and limits of agreement from 3.9% underestimation to 7.4% overestimation (Table 1; more detailed results in Supplementary Tables S4 and S5.) Heart rates were overestimated on average by 0.8 to 2.7 bpm, with limits of agreement of from 15.4bpm underestimation to 16.9bpm overestimation.

**Table 1.** Results of Bland-Altman analysis (Bias and 95% limits of agreement) for each oximetry device measured at 2 minutes. The reference oximeter is denoted as “R”. In each case differences were calculated as the reference measurements (SpO_2_ from the reference oximeter, blood gas analysis, or manual heart rate) minus the value displayed on the test oximeter. Negative values therefore indicate overestimation by the test oximeter, and positive values indicate underestimation by the test oximeter.

| Device | Cost | Bias (95% LOA) for SpO<sub>2</sub> compared to reference device | Bias (95% LOA) for SpO<sub>2</sub> compared to blood gas analysis | Bias (95% LOA) for heart rate compared to manual pulse |
| --- | --- | --- | --- | --- |
| D1 | £49 | -2.4% (-7.4 to 2.6) | -0.5% (-5.8 to 4.8) | -1.9bpm (-12.6 to 8.7) |
| D2 | £229 | -1.6% (-7.1 to 3.9) | 0.7% (-4.9 to 6.4) | -2.2bpm (-15.6 to 11.3) |
| D3 | £284.96 | -0.6% (-4.9 to 3.6) | 1.4% (-3.5 to 6.2) | -0.9bpm (-12.9 to 11.0) |
| D4 | £69.50 | -2.6% (-6.5 to 1.3) | -1.2% (-5.0 to 2.6) | -1.9bpm (-13.9 to 10.1) |
| D5 | £37.50 | -1.1% (-5.8 to 3.6) | 0.8% (-4.8 to 6.3) | -0.8bpm (-16.9 to 15.4) |
| D6 | £22.95 | -2.4% (-7.4 to 2.7) | -0.6% (-5.9 to 4.8) | -2.7bpm (-14.8 to 9.5) |
| R | N/A | N/A | 2.3 % (-4.1 to 8.6) | -1.6bpm (-12.5 to 9.3) |

LOA: Limits of agreement

A post-hoc sensitivity analysis was carried out excluding oximetry readings from one home oxygen clinic patient with a blood gas result of 46.7%, which is a level where pulse oximeters are not expected to be well-calibrated. This analysis (Supplementary Table S6) resulted in narrower limits of agreement for all devices when compared to the reference device.

### Secondary analyses

In patients with a recorded blood gas result, biases for the test oximeters compared to blood gas analysis ranged from 1.4% underestimation to 1.2% overestimation (Table 1; more detailed results in Supplementary Tables S4 and S5.) The reference oximeter showed a bias of 2.3% underestimation compared to blood gases, with limits of agreement ranging from 8.6% underestimation to 4.1% overestimation.

There was no clear association between device cost and accuracy (Figure 2) comparing against the reference oximeter or against blood gas oxygen saturation. The average time taken to obtain a good quality value varied from 5s to 10s in the fingertip pulse oximeters, and 14s in the reference oximeter (Table 2).

**Table 2.** Average time to first good quality measurement, and results of Bland-Altman analysis (Bias and 95% limits of agreement) for each oximetry device after 30 seconds. The reference oximeter is denoted as “R”. In each case differences were calculated as the references (SpO_2_ from the reference oximeter, or manual heart rate) minus the value displayed on the test oximeter. Negative values therefore indicate overestimation by the test oximeter, and positive values indicate underestimation by the test oximeter.

| Device | Cost | Time to first good quality SpO<sub>2</sub> value<br>Median (IQR) | Bias (95% LOA) for SpO<sub>2</sub> compared to reference device | Bias (95% LOA) for heart rate compared to manual pulse |
| --- | --- | --- | --- | --- |
| D1 | £49 | 5.0s (4.0 to 8.1) | -2.6% (-7.6 to 2.4) | -0.3bpm (-13.1 to 12.5) |
| D2 | £229 | 5.0s (4.2 to 6.5) | -1.3% (-7.5 to 4.8) | -2.1bpm (-17.4 to 13.2) |
| D3 | £284.96 | 10.0s (8.0 to 11.5) | -0.3% (-4.4 to 3.7) | -0.1bpm (-20.3 to 20.1) |
| D4 | £69.50 | 5.0s (4.0 to 5.5) | -2.0% (-4.5 to 0.6) | -1.9bpm (-11.4 to 7.6) |
| D5 | £37.50 | 8.5s (7.0 to 10.4) | -0.9% (-6.2 to 4.4) | -1.3bpm (-16.8 to 14.2) |
| D6 | £22.95 | 8.5s (7.0 to 10.5) | -2.3% (-8.2 to 3.6) | -1.8bpm (-11.4 to 7.8) |
| R |  | 14.0s (13.5 to 14.5) |  | -1.6bpm (-18.7 to 15.5) |

**Figure 2.**
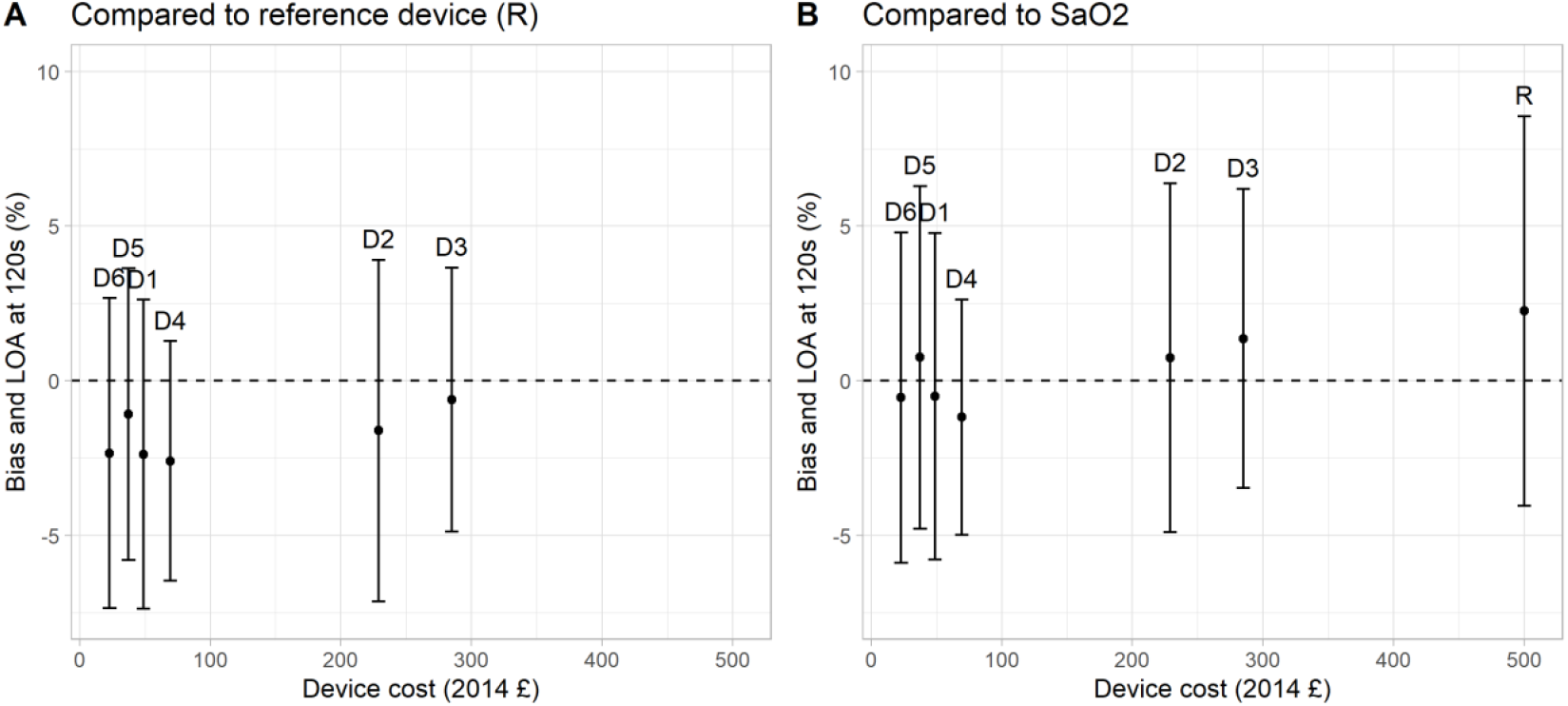
Accuracy of each device at 120 seconds (bias and limits of agreement from Bland-Alman analysis) by device purchase price. The reference device is given a nominal cost of £500 to allow for comparison.

Figure 3A demonstrates that all devices required a “settling time”, with values converging towards the final value at two minutes. The variability is also seen to decrease over time in Figure 3B. Most discrepancy from the final value and variability occur in the first 30 seconds of measurement (Figure 3, raw data presented in Supplementary Table S7). Table 2 shows the accuracy of the different oximeters at 30 seconds (detailed results in Supplementary Tables S5 and S8).

**Figure 3.**
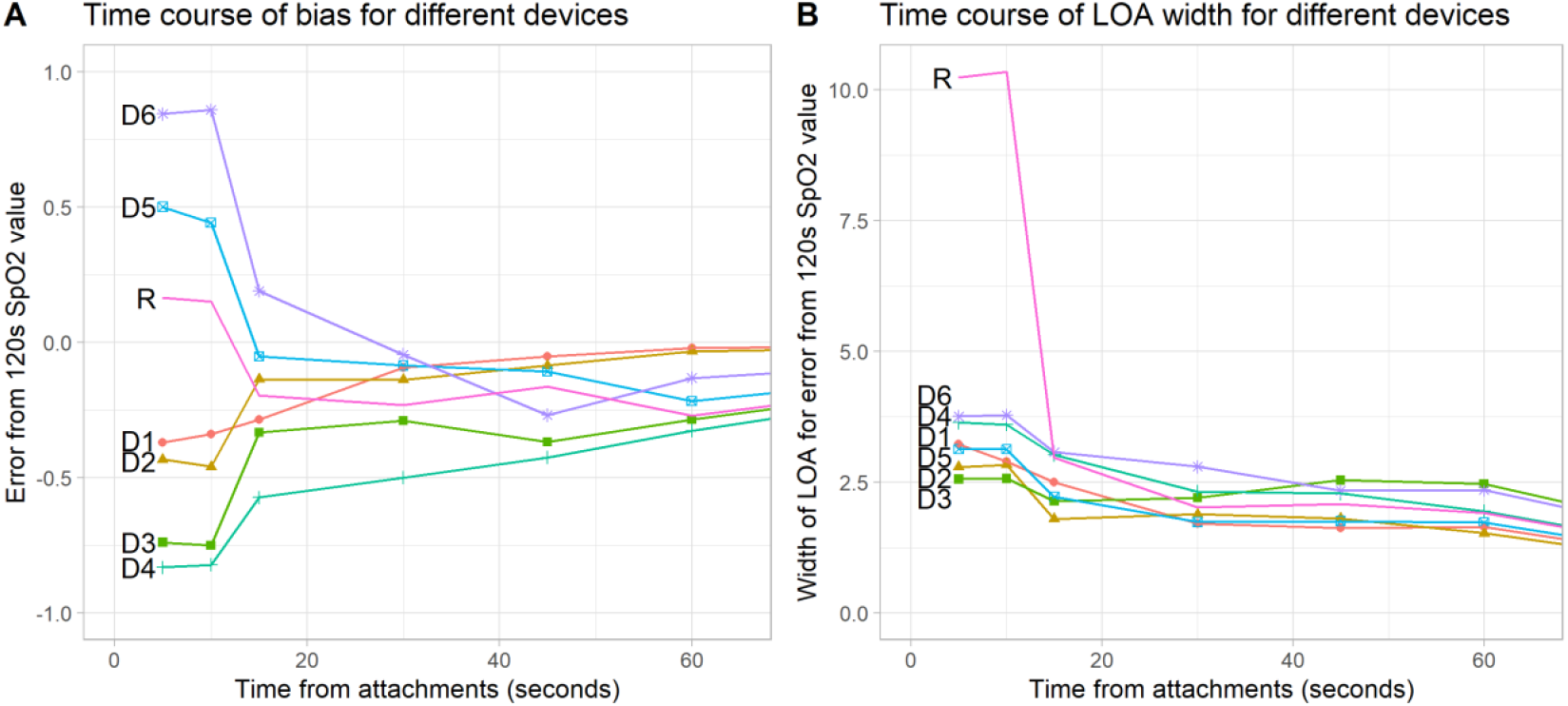
A) Time course of bias (average error) in device SpO_2_ measurements compared to measurement for that device at 2 minutes. B) Time course of LOA width (difference between the bias and the upper limit of agreement) of device SpO_2_ measurements In both plots, the first measurement is plotted at 5 seconds, and the first good quality measurement is plotted at 10 seconds, although these were measured at varying times.

There was no relationship between the time taken to display a good quality value, and the SpO_2_ displayed at two minutes (P=0.678 unadjusted; P=0.260 adjusting for device type; Supplementary Table S9).

## DISCUSSION

### Summary

The accuracy in commercially available fingertip pulse oximeters was variable, with average biases ranging from 0.6% to 2.6% overestimation, when compared to the reference oximeter. When compared to manual pulse measurement, oximeters tended to slightly overestimate heart rate, with biases ranging from 0.8 to 2.7 bpm. We observed the clinically noted phenomenon of “settling time” in both test and reference oximeters, occurring for some time after attaining a “good quality” trace, with readings trending towards the reading at 2 minutes. We did not identify any clear association between device cost and accuracy.

### Strengths and limitations

We tested a variety of pulse oximeters across the range of those in use by community health practitioners and patients against a recognised “gold standard” pulse oximeter used in current respiratory medicine practice to make clinical decisions. Study subjects were either healthy, or were respiratory patients with conditions affecting oxygen saturations, giving assessment of accuracy across a range of oxygen saturation values. We tested people at rest and in a stable state of health to ensure consistent oxygen levels, however our overall findings should be generalisable to less stable patients presenting more unwell.

This was a pragmatic study in a real-life clinical setting, with comparison to blood gas measurement where available. Data on the type of blood gas measurement (capillary or arterial) was not available so it was not possible to differentiate between these methods in the analysis. Capillary blood gas measurement may underestimate oxygen saturation compared to arterial measurements so this may have affected comparison of blood gas SaO_2_ with oximeter SpO_2_.

It is recognised that the costs and oximeters in this study date from 2014. However, pulse oximetry technology has not changed considerably, and all of the oximeter models tested in this study are still available for sale in 2024. We therefore expect that a study carried out now would have very similar findings.

### Comparison with existing literature

The biases and limits of agreement seen in this study are comparable to other studies in clinical settings assessing “cheap” widely available oximeters, although this is the first to assess changes in accuracy at different time points from application.(15,16,20,21)

An increasing number of studies have reported reduced pulse oximeter accuracy in people with darker skin tones, and this was particularly of concern during the COVID-19 pandemic where this risked underestimating severity of disease. (9,22–26) Most participants in this study were assessed as having “light” skin colour, so we have not attempted to carry out a subgroup analysis by skin colour as this would be underpowered.(16,21)

### Implications for research and/or practice

SpO_2_ measurements made with fingertip pulse oximeters may overestimate by up to 2.5% on average, which for most purposes would not be highly clinically significant. However errors exceeded 7% in around 1 in 20 measurements, which could have considerable impact on the clinical management of a patient (e.g. lack of escalation in an acutely unwell patient due to a falsely reassuring result, or changes in supplemental oxygen administration). Clinicians should consider blood gas measurement where higher accuracy is required, or consider repeating the measurement with an alternative device if there is uncertainty or poor correlation with other clinical findings.

Heart rates measured by test pulse oximeters were comparable to that measured by palpation of the radial artery, and were usually accurate to within 10 bpm, meaning that heart rate readings from pulse oximeters can typically be relied upon in clinical practice. We did not test accuracy of pulse oximeter heart rates in patients with arrythmias such as AF, and accuracy may be reduced in these patients.

Our results confirm that pulse oximeters require a period of time to display an accurate “good quality” SpO_2_ measurement, followed by a further period of “settling time” to attain a final reading. The first measure displayed on the screen should not be used for clinical decision making; we recommend waiting for at least 30 seconds from oximeter application to measurement recording to ensure sufficient accuracy.

We did not find a clear relationship between oximeter accuracy and cost, therefore this cannot be used to identify devices that will be more accurate in clinical practice.

We only recruited adults for this study, so the results may not apply to children, particularly younger children where attaching the device and keeping the limb still can be more problematic. Motion artefact is recognised as a potential cause of inaccuracy during pulse oximetry,(27) and was not assessed in this study.

### Conclusion

Fingertip pulse oximeters tended to overestimate SpO_2_ by up to 2.5%, but errors may exceed 7% in around 1 in 20 measurements, and operators should wait at least 30 seconds from oximeter application before reading the result. Price does not necessarily correlate with accuracy when considering purchasing a pulse oximeter. Heart rate was correct to within 10 seconds compared to manual pulse measurement.

## Supporting information

protocol

STARD

## Data Availability

The data that support the findings of this study are available from the corresponding author upon reasonable request.

## FUNDING

The study was funded by the Royal College of General Practitioners Scientific Foundation Board, the National Institute for Health and Care Research (NIHR) Community Healthcare MedTech and In Vitro Diagnostics Co-operative (MIC) and the NIHR HealthTech Research Centre (HRC) in Community Healthcare at Oxford Health NHS Foundation Trust. The views expressed are those of the authors and not necessarily those of the NHS, the NIHR or the Department of Health and Social Care. The funders had no input into the design, conduct, or reporting of the study.

## ACKNOWLEDGEMENTS

We are grateful to the staff and patients of the Home Oxygen Clinic at the Churchill Hospital for their patience and willingness to participate in the study. Particular thanks to Jan Turner-Wilson for support in designing the study and collecting data.

## COMPETING INTERESTS

The authors have no conflicts of interest to declare.

## ETHICAL APPROVAL

Ethic approval for the study was obtained from the NRES Committee East Midlands – Leicester (REC reference 14/EM/1098). The study sponsor was the University of Oxford.

## PATIENT AND PUBLIC INVOLVEMENT

Patients and the public were not involved in the development or conduct of the study. However, we have created a lay summary of the research results and intend to disseminate this to relevant patient groups after publication.

## AUTHOR CONTRIBUTIONS

SF and NMR designed the study. SF assisted with data collection. SF and MT extracted data from video recordings. SF was a major contributor to writing the manuscript. MT, NMR and HFA contributed to the manuscript, and read and approved the final version.

## Notes

### Competing Interest Statement

The authors have declared no competing interest.

### Author Declarations

NRES Committee East Midlands Leicester of NHS gave ethical approval for this work

