## Supplementary material for "Accuracy of fingertip pulse oximeters: a device accuracy study": protocol

**Study Title: Assessment of the accuracy of commercial pulse oximeters**

**Ethics Ref:** TBC

**EudraCT Number**: TBC

**Date and Version No**: 12 January 2015, version 1.6

| | |
|---|---|
| **Chief Investigator:** | Susannah Fleming<br>Nuffield Department of Primary Care Health Sciences<br><br>01865 289220 |
| **Investigators:** | Matthew Thompson (Nuffield Department of Primary Care Health Sciences)<br>Najib Rahman (Oxford Centre for Respiratory Medicine)<br>Jan Turner-Wilson (Oxford Centre for Respiratory Medicine) |
| **Sponsor:** | University of Oxford<br>Joint Research Office<br>Block 60<br>Churchill Hospital<br>Old Road<br>Headington<br>Oxford OX3 7LE |
| **Funder:** | Royal College of General Practitioners<br>Clinical Innovation and Research Centre<br>30 Euston Square<br>London NW1 2FB |
| **Chief Investigator Signature:** | |

All investigators declare that there are no actual or potential conflicts of interest.

**Confidentiality Statement**

This document contains confidential information that must not be disclosed to anyone other than the Sponsor, the Investigator Team, host organisation, and members of the Research Ethics Committee, unless authorised to do so.

© Copyright: The University of Oxford and Oxford University Hospitals NHS Trust 2014

© Copyright: The University of Oxford and Oxford University Hospitals NHS Trust 2014

10.4. Expectedness ... 18
10.5. Safety Monitoring ... 18
11. STATISTICS ... 18
11.1. Extraction of data from video recordings ... 18
11.2. Database entry ... 18
11.3. Inter-observer reliability of video extraction ... 18
11.4. Statistical analysis ... 19
Interim analysis ... 19
Full analysis ... 19
11.5. The Number of Participants ... 20
11.6. Procedure for Accounting for Missing, Unused, and Spurious Data. ... 20
11.7. Inclusion in Analysis ... 20
11.8. Procedures for Reporting any Deviation(s) from the Original Statistical Plan ... 20
12. DATA MANAGEMENT ... 21
12.1. Source Data ... 21
12.2. Access to Data ... 22
12.3. Data Recording and Record Keeping ... 22
13. QUALITY ASSURANCE PROCEDURES ... 22
14. SERIOUS BREACHES ... **Error! Bookmark not defined.**
15. ETHICAL AND REGULATORY CONSIDERATIONS ... 23
15.1. Declaration of Helsinki ... 23
15.2. ICH Guidelines for Good Clinical Practice ... 23
15.3. Medical Device regulations ... **Error! Bookmark not defined.**
15.4. Approvals ... 23
15.5. Reporting ... 23
15.6. Participant Confidentiality ... 23
15.7. Other Ethical Considerations ... 23
16. FINANCE AND INSURANCE ... 24
16.1. Funding ... 24
16.2. Insurance ... 24
17. PUBLICATION POLICY ... 24
Authorship ... 24
Acknowledgements ... 24
Informing participants ... 25
18. REFERENCES ... 25

© Copyright: The University of Oxford and Oxford University Hospitals NHS Trust 2014

19. APPENDIX A: AMENDMENT HISTORY ........ 26
20. Appendix B: Equation for 95% CI of sample standard deviation ........ 26

#### 1. KEY STUDY CONTACTS

| | |
|---|---|
| **Chief Investigator** | Susannah Fleming<br>Nuffield Department of Primary Care Health Sciences<br>Radcliffe Observatory Quarter<br>Woodstock Road<br>Oxford OX2 6GG<br>phone: 01865 289220<br> |
| **Sponsor** | University of Oxford<br>Joint Research Office<br>Block 60<br>Churchill Hospital<br>Old Road<br>Headington<br>Oxford OX3 7LE<br>phone: 01865 572221<br><br>Fax: 01865 572228 |
| **Clinical Lead and Principal Investigator at NHS Site** | Dr Najib Rahman<br>Oxford Centre for Respiratory Medicine<br>Churchill Hospital<br>Old Road<br>Headington<br>Oxford OX3 7LE<br>phone: 01865 225230<br><br>fax: 01865 225221 |

#### 2. SYNOPSIS

| | |
|---|---|
| Study Title | Assessment of the accuracy of commercial pulse oximeters |
| Study Design | Device accuracy study |
| Study Participants | Patients attending home oxygen clinic and healthy adult volunteers |
| Planned Sample Size | 75 |
| Treatment duration | Single study visit – no treatment above standard care planned |

© Copyright: The University of Oxford and Oxford University Hospitals NHS Trust 2014

| Planned Study Period | August 2014 – May 2015 | |
|---|---|---|
| | Objectives | Outcome Measures/Endpoints |
| Primary | Do commercially available pulse oximeters report the correct oxygen saturation value in patients with a variety of clinically relevant oxygen levels? | **1) Difference between oxygen saturations reported by index and reference oximeters at 120 seconds from application (primary outcome)**<br>2) Difference between oxygen saturations reported by pulse oximeters at 120 seconds from application and blood gas measurements (where measured)<br>3) Difference between oxygen saturations reported by index oximeters at 15, 30, 45, and 60 seconds from application, and reference oximeter value at 120 seconds from application<br>4) Difference between oxygen saturations reported by pulse oximeters at 15, 30, 45, and 60 seconds from application and blood gas measurements (where measured) |
| Secondary | 1) Do commercially available pulse oximeters report the correct pulse rate in patients with a variety of clinically relevant oxygen levels?<br>2) How long do commercially available pulse oximeters take to report a good quality oxygen saturation value in patients with a variety of clinically relevant oxygen levels?<br>3) Do some commercially available pulse oximeters show a delay ("settling time") between the first oxygen saturation value reported, and reporting the true oxygen saturation value? | 1) Difference between heart rate reported by pulse oximeters at 120 seconds from application and radial pulse measurement<br>2) Difference between heart rate reported by pulse oximeters at 15, 30, 45, and 60 seconds from application and radial pulse measurement<br>3) Time between application and first "good quality" value for each oximeter<br>4) Time between application and first reported oxygen saturation value for each oximeter<br>5) Difference in oxygen saturation for each oximeter between first reported value and value reported at 120s from application<br>6) Difference in oxygen saturation for each oximeter between first "good quality" value and value reported at 120s from application |
| Device names | Merlin M-Pulse Lite, Nonin Onyx II 9550, BCI 3301, Choicemed MD300, Contec CMS-50D, Contec CMS-50DL. Ohmeda 3700 (reference) | |
| Device Manufacturers | Merlin medical, Nonin, BCI, ChoiceMMed, Contec Medical Systems, Ohmeda | |

© Copyright: The University of Oxford and Oxford University Hospitals NHS Trust 2014

| | |
|---|---|
| Device Classification | CE-marked pulse oximeters on general sale |

#### 3. ABBREVIATIONS

| | |
|---|---|
| AE | Adverse event |
| AR | Adverse reaction |
| CI | Chief Investigator |
| CRA | Clinical Research Associate (Monitor) |
| CRF | Case Report Form |
| CRO | Contract Research Organisation |
| CT | Clinical Trials |
| CTA | Clinical Trials Authorisation |
| CTRG | Clinical Trials and Research Governance |
| DMC/DMSC | Data Monitoring Committee / Data Monitoring and Safety Committee |
| DSUR | Development Safety Update Report |
| GCP | Good Clinical Practice |
| GP | General Practitioner |
| GTAC | Gene Therapy Advisory Committee |
| IB | Investigators Brochure |
| ICF | Informed Consent Form |
| ICH | International Conference of Harmonisation |
| IMP | Investigational Medicinal Product |
| IRB | Independent Review Board |
| MHRA | Medicines and Healthcare products Regulatory Agency |
| NHS | National Health Service |
| NRES | National Research Ethics Service |
| OXTREC | Oxford Tropical Research Ethics Committee |
| PI | Principal Investigator |
| PIL | Participant/ Patient Information Leaflet |
| R&D | NHS Trust R&D Department |
| REC | Research Ethics Committee |
| SAE | Serious Adverse Event |
| SaO2 | Oxygen saturation measured by blood gas measurement |
| SAR | Serious Adverse Reaction |
| SDV | Source Data Verification |

© Copyright: The University of Oxford and Oxford University Hospitals NHS Trust 2014

| | |
|---|---|
| SMPC | Summary of Medicinal Product Characteristics |
| SOP | Standard Operating Procedure |
| SpO2 | Oxygen saturation measured by pulse oximetry |
| SUSAR | Suspected Unexpected Serious Adverse Reactions |
| TMF | Trial Master File |
| TSG | Oxford University Hospitals Trust / University of Oxford Trials Safety Group |

### 4. BACKGROUND AND RATIONALE

#### 4.1. Overview

Pulse oximeters are used widely by a variety of health professionals, including general practitioners. A wide variety of makes and models of pulse oximeter are available, ranging in price from around £20 to over £1000. There is very little information available on the accuracy of these devices, particularly at the cheaper end of the market, and some evidence that not all devices are capable of picking up clinically important drops in oxygen levels.[1,2]

We wish to test the accuracy of a variety of commercially available pulse oximeters on human volunteers with a range of clinically relevant oxygen levels.

We know from a survey of general practitioners and practice nurses that many cheap pulse oximeters are in use in general practice, and that some clinicians have concerns about their ability to detect patients with low blood oxygen levels. Measurements made by pulse oximeters are used to assess the severity of various conditions, particularly respiratory conditions, and to determine whether additional oxygen therapy is required. Incorrect readings, particularly incorrectly high readings, may result in false reassurance or inappropriate therapy decisions. This may lead to delays in diagnosis or treatment, and increased morbidity.

#### 4.2. Literature review

A literature search identified few studies comparing the accuracy of different modern pulse oximeter models, all of which were focussed on models used in hospitals, rather than those used in primary care, which are likely to be cheaper, due to the lesser utilisation of oximetry in primary care, and the greater need for one-off "spot checks" rather than continuous monitoring.

Milner *et al* [1] used a calibration tester to assess 847 oximeters in clinical use in 29 NHS hospitals. This identified that 30.5% of the oximeters were not operating in accordance with the manufacturer's specifications. In an *in vivo* study, Feiner *et al* [3] monitored 36 healthy subjects using both blood gas measurement and three commercial hospital-grade pulse oximeters during inspiration of hypoxic gas mixtures. This study found large biases at extremely low levels of oxygen saturation (<80%), but not at levels more commonly found in clinical practice. Other studies confirmed known limitations of pulse oximetry in patients with particular conditions (e.g. sickle cell anaemia, preterm infants, carbon monoxide poisoning), but did not investigate the accuracy of cheaper pulse oximeters in patients exhibiting oxygen saturation levels that might be expected to be seen in general practice.

© Copyright: The University of Oxford and Oxford University Hospitals NHS Trust 2014

##### 4.3. Summary of research plan

We intend to test the accuracy of six index pulse oximeters, which are known to be used in general practice. Accuracy of oxygen saturation readings will be compared to those obtained from a reference pulse oximeter (pre-calibrated or FDA-approved), and blood gas measurements, which will be taken on participants who require blood gas measurement for clinical management. Accuracy of heart rate measurements will be compared to manually-measured radial pulse rates.

In addition, we will look for evidence of the anecdotally-observed "settling time" in pulse oximeters, where the initial reading is different to that displayed after a period of monitoring.

As it is important to assess the accuracy of pulse oximeters over a range of physiological levels of oxygen saturation, we will recruit participants from two populations. To obtain readings corresponding to normal physiological levels of oxygen saturation (96-100%), we will recruit healthy adults, and to obtain reading corresponding to lower levels, we will recruit patients from a respiratory outpatients clinic with a high proportion of patients with chronically low oxygen saturations (<96%).

To ensure accurate assessment of the readings of multiple pulse oximeters, video recording will be used to record the displays of the pulse oximeters during applications, for later analysis.

#### 5. OBJECTIVES AND OUTCOME MEASURES/ENDPOINTS

| Objectives | Outcome Measures/Endpoints |
|---|---|
| **Primary Objective**<br>Do commercially available pulse oximeters report the correct oxygen saturation value in patients with a variety of clinically relevant oxygen levels? | **1) Difference between oxygen saturations reported by index and reference oximeters at 120 seconds from application (primary outcome)**<br>2) Difference between oxygen saturations reported by pulse oximeters at 120 seconds from application and blood gas measurements (where measured)<br>3) Difference between oxygen saturations reported by index oximeters at 15, 30, 45, and 60 seconds from application, and reference oximeter value at 120 seconds from application<br>4) Difference between oxygen saturations reported by pulse oximeters at 15, 30, 45, and 60 seconds from application and blood gas measurements (where measured) |
| **Secondary Objectives**<br>1) Do commercially available pulse oximeters report the correct pulse rate in patients with a variety of clinically relevant oxygen levels? | 1) Difference between heart rate reported by pulse oximeters at 120 seconds from application and radial pulse measurement<br>2) Difference between heart rate reported by pulse oximeters at 15, 30, 45, and 60 seconds from application and radial pulse measurement |
| **Secondary Objectives**<br>2) How long do commercially available pulse | 1) Time between application and first "good quality" value for each oximeter |

© Copyright: The University of Oxford and Oxford University Hospitals NHS Trust 2014

| | |
|---|---|
| oximeters take to report a good quality oxygen saturation value in patients with a variety of clinically relevant oxygen levels? | 2) Time between application and first reported oxygen saturation value for each oximeter |
| **Secondary Objectives**<br>3) Do some commercially available pulse oximeters show a delay ("settling time") between the first oxygen saturation value reported, and reporting the true oxygen saturation value? | 1) Difference in oxygen saturation for each oximeter between first reported value and value reported at 120s from application<br>2) Difference in oxygen saturation for each oximeter between first "good quality" value and value reported at 120s from application |

#### 6. STUDY DESIGN

This is primarily a method comparison study, comparing the measurements made by index pulse oximeters to those made by a reference oximeter, and to "gold standards" of blood gas measurement (where available), and manual radial pulse rate. Figure 1 shows participant flow through the different study procedures.

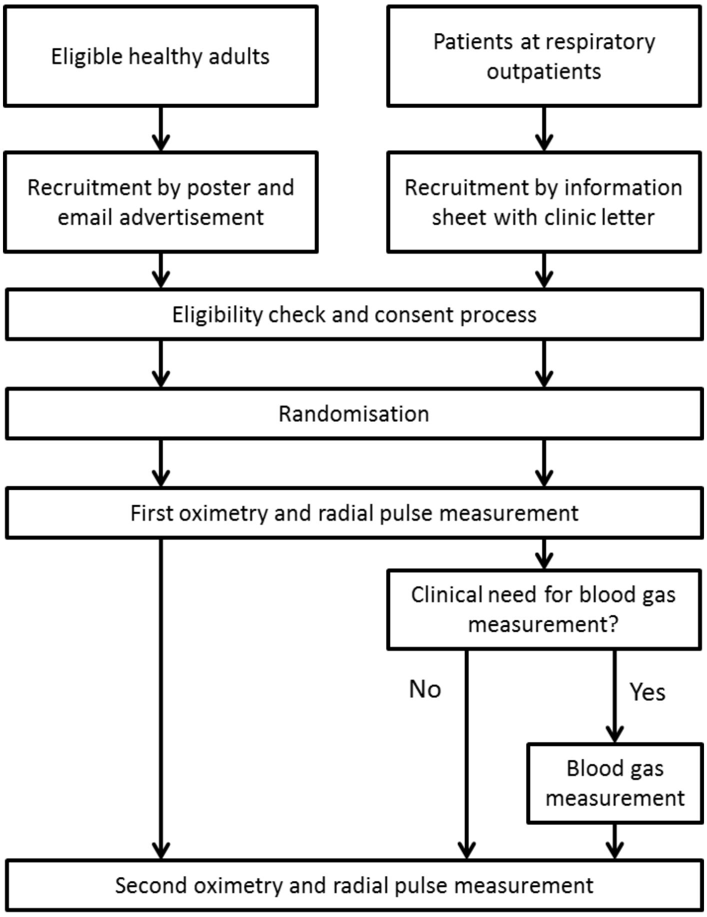

**Figure 1: Schematic diagram of study procedures**

The entire study will take place in a single visit at the respiratory outpatient department in the Churchill hospital. The eligibility check and consent procedure are likely to take 15-30 minutes per participant, with shorter times expected for healthy participants.

© Copyright: The University of Oxford and Oxford University Hospitals NHS Trust 2014

Pulse oximetry will be carried out with 4 pulse oximeters (3 index and 1 reference device) on the four fingers of the right hand, with the displays of the pulse oximeters being videoed for 2 minutes. During this time, a manual radial pulse measurement will be taken. A second measurement will then be carried out with the remaining 3 index devices and the reference device for 2 minutes, again with a radial pulse measurement. Devices will be randomised to fingers and, in the case of index devices, to the first or second measurement. If clinically indicated, a blood gas measurement will be taken between the two oximetry measurements.

The estimated total time required for both oximetry measurements is 30-45 minutes per participant, with shorter times expected for healthy participants, and those who do not require blood gas measurement.

#### 7. PARTICIPANT IDENTIFICATION

##### 7.1. Study Participants

75 participants will be recruited to the study. Of these, at least 20 should have consistent reference pulse oximetry readings of 96-100% (as recorded at the end of both 2-minute recording periods).
Participants will be either healthy volunteers, or patients attending the respiratory outpatients clinic.

##### 7.2. Inclusion Criteria

- Participant is willing and able to give informed consent for participation in the study.
- Male or Female, aged 18 years or above.
- Fulfills one of the following two criteria:
  - current patient due to attend the respiratory outpatients clinic during the study period
  - adult who is in good general health.

##### 7.3. Exclusion Criteria

The participant may not enter the study if ANY of the following apply:

- Current application of dyes, false nails, nail polish, or other cosmetics (e.g. henna) on the fingers of the right hand.
- Amputation of any part of the right upper limb resulting in loss of one or more fingers on the right hand.
- Recent (<1 week) medical procedure involving the administration of intravenous dyes or contrast media.

#### 8. STUDY PROCEDURES

##### 8.1. Preparation

© Copyright: The University of Oxford and Oxford University Hospitals NHS Trust 2014

Batteries used to power the various pulse oximeters will be checked for adequate voltage every 2 weeks using a commercial battery tester. Any batteries identified as weak or poor will be replaced.
All oximeters will be cleaned with Clinell wipes (or equivalent) and allowed to air dry between patients, following standard infection prevention and control procedures in place at the study site.
Randomisation will be carried out prior to recruitment, with randomisations secured in opaque envelopes labelled with the sequential study numbers.

##### 8.2. Recruitment

An initial cohort of 50 participants will be recruited from patients booked to attend the respiratory outpatients clinic. Details of the study, including a participant information sheet, will be sent out to potentially eligible (>18 years) patients with their appointment letter. Insertion of the study information will be carried out by NHS administrative personnel who would usually prepare patient invitation letters.
At least 20 participants are required with oxygen saturation levels of 96-100%. Such levels are rare in the population attending the respiratory outpatients clinic, and so additional healthy participants will need to be recruited to achieve this goal after oxygen saturation levels of the initial 50 participants are known. Healthy adults will be recruited via poster and email advertisements. Prospective participants will contact the study team, who will provide them with a participant information sheet at the earliest possible opportunity. Further recruitment (if required) up to the total sample size of 75 will take place in the respiratory outpatients clinic. For example, if 15 healthy participants are needed, a further 10 patients will be recruited to make a total sample size of 75 (50 initial cohort (including 5 with high saturations) + 15 healthy participants + 10 final cohort).

##### 8.3. Eligibility and Informed Consent

Eligibility criteria (defined above) will be clearly outlined in the participant information sheets sent out to potential participants. Prior to consent, eligibility will be confirmed by the researcher taking consent.

Written consent will be obtained from all participants prior to taking part in the study, and will be witnessed by a researcher. Participants will have the opportunity to ask questions about the study during the consent conversation, which will include written copies of the information sheet, and will cover the nature of the study, and what it will involve for the participant. It will be clearly stated that the participant is free to withdraw from the study at any time for any reason without prejudice to future care, and with no obligation to give the reason for withdrawal.

Participants whose capacity to give informed consent is impaired (e.g. by cognitive disability or intoxication) will not be permitted to take part in the study. In the unlikely event that a participant's capacity becomes impaired during the course of the study, they will be withdrawn and all readings discarded, as the short period of time between consent and the end of the study would render any previous consent suspect in such a circumstance.

The participant must personally sign and date the latest approved version of the Informed Consent form before any study measurements are performed.

After obtaining written consent for participation in the study (including videographic recording for the purposes of research), a similar process will be carried out to seek consent for publication of anonymised clips or stills from the videographic recordings taken during the course of the study. Consent for publication will be sought separately from consent for research, and it will be made clear that consent to publication is not a requirement for inclusion in the study.

© Copyright: The University of Oxford and Oxford University Hospitals NHS Trust 2014

The participant will be allowed as much time as wished to consider the information, and the opportunity to question the Investigator, their GP or other independent parties to decide whether they will participate in the study. Written Informed Consent will then be obtained by means of participant dated signature(s) and dated signature(s) of the person who presented and obtained the Informed Consent. The person who obtained the consent must be suitably qualified and experienced, and have been authorised to do so by the Chief/Principal Investigator. A copy of the signed Informed Consent form or forms will be given to the participant. The original signed forms will be retained at the study site.

##### 8.4. Randomisation, blinding and code-breaking

All participants will be identified by a study code number, assigned at the time of consent. Code numbers will be assigned in strictly consecutive order. Code numbers will be used on all study paperwork, including consent forms, case and video report forms, and randomisation paperwork. Participants' personal data will not appear on any form, with the exception of the consent form, which requires a printed name and signature.
There will be no procedure for deanonymising participants.

Seven models of pulse oximeter will be used in this study – one reference device (R), and six index devices (D1-D6). Two specimens of each model will be used, termed "a" and "b", to reduce the risk of spurious results due to a single malfunctioning oximeter. However, for logistical reasons, only one reference (R) device will be used. Thus, 13 devices in total will be used in the study. Each participant will be measured twice, with four oximeters used during each measurement. In each case, this will include one reference device, and three index devices. Over the two sets of measurements, each participant will be measured using each of the six index models.

Three levels of randomisation will be used when assigning individual devices to a participant.

The first level is randomisation of testing order. This determines which three of the six index models are tested in the first set of measurements, and which in the second.

Once the models in each measurement are determined, the exact device to be used is also randomised, between the "a" and "b" devices for each model.

Within each measurement, devices are randomised to fingers. This is to avoid any bias due to physiological differences, or delays between fitting devices to fingers (which is always carried out from index -> little finger). Thus a third randomisation is carried out to assign devices to the fingers of the participant's hand.

All randomisations will be carried out using random number sequences generated by random.org.

Randomisations for each participant will be placed in an opaque envelope labelled with the associated study code number. Distribution of randomisation envelopes will be carried out before recruitment of the first participant. Code numbers will be assigned in strict numerical order at the time of consent. Following consent, the corresponding randomisation envelope will be identified and opened to reveal the participant's randomisation.

***Example randomisation:***

*First level:*

R D2 D3 D5 in first measurement, R D1 D4 D6 in second measurement

*Second level:*

© Copyright: The University of Oxford and Oxford University Hospitals NHS Trust 2014

R D2b D3b D5a in first measurement, R D1b D4b D6a in second measurement

*Third level:*

First measurement:

Index: D3b

Middle: R

Ring: D5a

Little: D2b

Second measurement:

Index: R

Middle: D4b

Ring: D1b

Little: D6a

#### 8.5. Study measurements

##### First oximetry measurement

The subject will be seated comfortably, with the right hand resting on a table at or below the level of the heart. A video camera with an accurate internal clock will be set up to film the right hand of the participant. The camera will be set up such that the recording will not show the face or other identifying features, but will be able to record the displays of the pulse oximeters when they are attached to the fingers. The randomisation envelope will also be included in the view of the camera, to identify the recording, and the date and time will be recorded on the case form, as an additional method of matching video recordings to case forms.

The video recording will be started and the first four oximeters will be attached to the fingers of the right hand as specified in the randomisation, with the index finger device applied first, and proceeding medially. Oximeters will be attached as quickly as possible, while ensuring that they are securely and correctly placed on the fingers.

Once all four oximeters are attached, a two-minute timer will be started. While the timer is running, the heart rate of the participant will be assessed and recorded by manual counting of the radial pulse at the left wrist for one minute.

When two minutes have elapsed, the oximeters will be removed from the fingers.

##### Blood gas measurement

If participants have been recruited from respiratory outpatients, and a blood gas measurement is clinically indicated as part of their care, this will be taken between the first and second oximetry measurements. Normal clinic practice will be followed regarding the measurement procedure, and the result will be recorded on the case report form.

##### Second oximetry measurement

After the first oximetry measurement (and blood gas measurement, if taken) is complete, a second oximetry measurement will be made. The second four oximeters will be attached to the fingers of the right hand as specified in the randomisation, in the same manner as for the first measurement. A two minute timer will be started, and the radial pulse will be measured, as before.

© Copyright: The University of Oxford and Oxford University Hospitals NHS Trust 2014

When two minutes have elapsed, the oximeters will be removed from the fingers, and the video recording terminated.

##### 8.6. Discontinuation/Withdrawal of Participants from Study

Each participant has the right to withdraw from the study at any time. Where possible, withdrawn participants will be replaced to ensure an adequate sample size.

The reason for withdrawal will be recorded in the CRF.

If the participant is withdrawn due to an adverse event, the Investigator will arrange for follow-up visits or telephone calls until the adverse event has resolved or stabilised.

##### 8.7. Definition of End of Study

The end of stduy is the date of the last visit of the last participant.

#### 9. Identification & Description of the Investigational Device

Index devices were chosen based on the results of a survey of general practitioners and research nurses, so that they would be representative of devices in current use in general practice. A postal questionnaire was sent out to all general practitioners and research nurses recruiting participants to the "3C" study, whose records showed that they had recruited a participant with an oximetry reading since October 2011. Questionnaires were sent out to 833 clinicians, asking them to report the make and model of any oximeters that they personally used. To assist in the identification, photographs of 43 oximeters on sale to general practitioners were included in the questionnaire. Entrance into a draw for an iPod Nano was offered as an incentive for return of the questionnaire.

Of the 833 surveys sent out to clinicians, 558 (67%) were returned. Respondents reported using a wide variety of devices, including ones not included in the 43 photographed on the survey. Many respondents indicated that they used more than one device.

The two most commonly indicated devices, present on more than a third of returned surveys were the Merlin M-pulse (including the M-pulse lite model), which was indicated by 231 (41.4%) respondents, and the Nonin Onyx (including the Onyx II and Nonin Vantage 9500, 9550 and 9560 models), indicated by 202 (36.2%) respondents.

Three further "devices" were indicated by more than 2% of respondents. The BCI 3301 was indicated by 38 respondents (6.8%). In the free text response to using a device other to those shown on the survey, two manufacturer names appeared on more than 2% of the total number of surveys. Since further research has indicated that model names for these manufacturers are difficult to interpret and may be very similar in function, they have been grouped together as single devices. These two "devices" are Choicemed, indicated in the "other" category by 23 respondents (4.1%), but also present as a device on the survey, increasing this to 26 respondents (4.4%), and Contec, indicated in the "other" category by 21 respondents (3.8%).

© Copyright: The University of Oxford and Oxford University Hospitals NHS Trust 2014

A further 2 devices appear in 10 or more responses. "Biosync" devices were cited in the "other" category by 12 respondents (2.0%), and Daray V408/V409 were indicated by 10 respondents (1.8%).

All other devices or manufacturers (in the case of responses in the "other" category) appeared in less than 10 responses.

#### 9.1. Device description

The devices to be used in this study are CE-marked pulse oximeters available for general sale. These are used for one-off and ongoing monitoring of heart rate and oxygen saturation (SpO2) in a variety of clinical settings. Within the context of this study, the devices will be used for their intended purpose, as the study is intended to assess the accuracy of various models and makes of devices on sale and used by general practitioners.

No specific training is required to operate a pulse oximeter, and they are frequently used by healthcare assistants within clinical settings. However, a standard operating procedure for their use within the study has been developed, and will be followed by the research nurses who will be using the oximeters. Research nurses will be familiar with respiratory research, and will therefore be well versed in obtaining pulse oximetry measurements.

##### Reference device

The reference pulse oximeter will be a pulse oximeter that has been pre-calibrated by the hospital medical physics department, or if this is not possible, an oximeter which is known to be FDA-approved, and which is used with Oxford University Hospital Trust for in-patient monitoring.

##### Index devices

Six index device models will be used in the study (listed below). These were identified based on the most popular devices reported by respondents to the postal survey. All devices are CE-marked.

1. Merlin M-pulse Lite
2. Nonin Onyx II 9550
3. BCI 3301
4. Choicemed MD300
5. Contec CMS-50D
6. Contec CMS-50DL

Two devices from each model will be used, denoted 'a' and 'b'. A log of serial numbers for both 'a' and 'b' devices (and in the case of the BCI 3301), the detachable finger probes will be kept. Each participant will be randomised to either an 'a' or 'b' device from each pair, which will be noted on the CRF, allowing traceability.

#### 9.2. Device Safety

All devices will be purchased new from commercial suppliers, and will be used in line with the manufacturers' instructions. All devices will be inspected for CE marking prior to use, and will be battery

© Copyright: The University of Oxford and Oxford University Hospitals NHS Trust 2014

powered. The exception to this is the reference device, which may be CE marked and mains-powered, provided that it displays evidence of having been tested for electrical safety.

##### 9.3. Device Accountability

All devices will be stored in a secure indoor location, free from excessive temperature or humidity.

#### 10. SAFETY REPORTING

##### 10.1. Definitions

<table>
<tr><td>Adverse Event (AE)</td><td>Any untoward medical occurrence in a participant to whom a medicinal product has been administered, including occurrences which are not necessarily caused by or related to that product.</td></tr>
<tr><td>Adverse Device effect (ADE)</td><td>An adverse event related to the use of an investigational medical device. This definition includes any events resulting from insufficient or inadequate instructions for use, deployment, implantation, installation, or operation, or any malfunction of the investigational device. This definition also includes any event resulting from user error or form intentional misuse of the investigational device.</td></tr>
<tr><td>Serious Adverse Event (SAE)</td><td>An adverse event that:<br>• Led to death<br>• Resulted in serious deterioration in the health of the subject that:<br>o resulted in a life-threatening illness or injury<br>o resulted in a permanent impairment of a body structure or a body function<br>o required in-patient care or prolongation of hospitalisation<br>o resulted in medical or surgical intervention to prevent life-threatening illness or injury or permanent impairment to a body structure or a body function.<br>This includes device deficiencies that might have led to a serious adverse event if:<br>a) suitable action had not been taken or<br>b) intervention had not been made or<br>c) circumstances had been less fortunate.<br>These are handled under the SAE reporting system.<br>Planned hospitalisation for a pre-existing condition, or a procedure</td></tr>
</table>

© Copyright: The University of Oxford and Oxford University Hospitals NHS Trust 2014

| | |
|---|---|
| | required by the study protocol, without serious deterioration in health, is not considered a serious adverse event. |
| Serious Adverse Device Effect (SADE) | Any untoward medical occurrence that can be attributed wholly or partly to the device, which resulted in any of the characteristics of a serious adverse event as described above.<br><br>*Unanticipated Serious Adverse Device Effects (USADE)*<br><br>Any serious adverse device effect which, by its nature, incidence, severity or outcome, has not been identified |
| Device deficiency | Inadequacy of a medical device with respect to its identity, quality, durability, reliability, safety or performance. Device deficiencies include malfunctions, use errors and inadequate labeling.<br><br>Device deficiencies that did not lead to an adverse event, but could have led to a medical occurrence if suitable action had not been taken, or intervention had not been made or if circumstances had been less fortunate |
| User error | Act or omission of an act that results in a different medical device response than intended by the manufacturer or expected by the user. Use error includes slips, lapses and mistakes. An unexpected physiological response of the subject does not itself constitute a use error. |

*Severity definitions*

The following definitions will be used to determine the severity rating for all adverse events:

Mild: awareness of signs or symptoms, that does not interfere with the subject's usual activity or is transient that resolved without treatment and with no sequelae.

Moderate: a sign or symptom, which interferes with the subject's usual activity.

Severe: incapacity with inability to do work or perform usual activities.

#### 10.2. Procedures for Recording Adverse Events

The safety record of pulse oximeters is very good (as evidenced by their deployment in a wide range of clinical environments and use by minimally trained personnel). We therefore believe that is appropriate to only report unexpected device-related serious adverse events in this study.

#### 10.3. Reporting Procedures for Serious Adverse Events

Reporting of all Serious Adverse Events will be done in accordance with the European Commission Guidelines on Medical Devices Serious Adverse Event Reporting (MEDDEV 2.7/3; December 2010).

© Copyright: The University of Oxford and Oxford University Hospitals NHS Trust 2014

All unexpected device-related SAEs will be reported to the manufacturer and the MHRA, using the online form on the MHRA website at :

http://www.mhra.gov.uk/Safetyinformation/Reportingsafetyproblems/Devices/index.htm

SAEs will be reported immediately or no later than 24 hours after the investigator becomes aware to the device manufacturer and regulatory authority using the reporting form at the site above. SAEs that occur within 24 hours of last use of the device will be recorded.

##### 10.4. Expectedness

Expectedness will be determined according to the Manufacturers Instruction Manual for the device.

##### 10.5. Safety Monitoring

As the devices are CE-marked and being used for their designed purposed, this study does not fall under the regulations and no safety monitoring committee will be convened.

#### 11. STATISTICS

##### 11.1. Extraction of data from video recordings

Following data recording, data will be extracted from the video recording in the following manner. Each video recording will be watched by two independent observers, who will record the following for each of the two oximetry measurements on a video reporting form:

- Time at which each device is placed on the finger (defined as when the researcher is no longer touching it)
- Time at which each device first displays oxygen saturation reading, and saturation reading displayed at this time
- Time at which each device first displays heart rate reading, and heart rate reading displayed at this time
- Time at which each device first displays “good quality” reading (judged according to manufacturers’ instructions e.g. by observing a pulse strength indicator), and both oxygen saturation and heart rate readings displayed at this time
- Heart rate and oxygen saturation readings on all devices at 15, 30, 45, 60 and 120 seconds from the time the last device is placed.

Readers will be blinded to both the results of blood gas and manual radial pulse measurements.

##### 11.2. Database entry

Data from the case report forms and video reporting forms will be entered onto a database using dual entry, with disagreements resolved by reference to the original forms, and adjudication by a third reader if necessary.

##### 11.3. Inter-observer reliability of video extraction

© Copyright: The University of Oxford and Oxford University Hospitals NHS Trust 2014

Video extraction will be carried out by two independent, blinded, observers, as described in the previous section. Inter-observer reliability measures will be calculated for the various measures recorded from the video. Since all of these measures are continuous rather than binary or categorical, inter-observer reliability will be assessed using the intra-class correlation coefficient.

#### 11.4. Statistical analysis

The main analysis of the final results of the study will be via Bland-Altman plots, which will provide an indication of the limits of agreement between the index test and reference standard. These plots will also allow identification of both systematic and proportional bias in the results.

##### Interim analysis

An interim analysis will be carried out after 50 participants have been recruited. It is anticipated that all of these participants will have been recruited from the respiratory outpatients clinic.

An assessment will be made of how many of the participants fulfil the criteria for having a consistent oxygen saturation level of 96-100%. This will be used to determine the recruitment split for the remaining participants between respiratory outpatients and healthy adults, with the intention that 20-25 of the final sample of 75 participants will have a consistent oxygen saturation level of 96-100%.

An additional check will also be made to ensure that the assumptions underlying the sample size calculation are valid. The sample size calculation is dependent on the standard deviation of the difference between oxygen saturation values (s). This was estimated as 2.5%. If this is an overestimate, then our sample size is valid, albeit overestimated. However, if this has been underestimated, then we may need a larger sample size to obtain the required 1% accuracy for the 95% confidence intervals of the limits of agreement on the Bland-Altman plots.

The sample standard deviations for each of the differences in the two primary endpoints will therefore be calculated, along with their 95% confidence intervals (equation given in Appendix B). If the lower 95% confidence interval is less than 2.5% for all samples, we can assume that our initial estimate was reasonable, or an overestimate, and so our sample size is acceptable. However, if the lower bound of the 95% confidence interval is greater than 2.5% for any of the samples, then our initial calculated sample size may be insufficient, and it may be necessary to increase the intended number of recruited subjects.

In this case, interim Bland-Altman plots for the primary endpoints will be produced to assist the investigators in deciding whether a larger sample size is required.

##### Full analysis

All collected data will be included in the analysis, with the exception of data from subsequently withdrawn participants. All efforts will be made to obtain a full and complete data set. Where data is unavoidably lost, it will not be imputed. However, in the case of incomplete data from a given participant, this will be included in all analyses for which full data is available. Since most analyses require only two data points, it is envisaged that occasional missing data will not have a significantly adverse effect on the results of the study.

Most of the endpoints in this study are differences in oxygen saturation, differences between oxygen saturation and blood gas measurement ($SaO_2$), and differences in heart rate. These will all be assessed using Bland-Altman plots, with logarithmic transformation and/or linear regression in cases where the

© Copyright: The University of Oxford and Oxford University Hospitals NHS Trust 2014

Bland-Altman plot shows indications of proportional bias (i.e. the difference is related to the mean value).

If any oximeters show clinically important differences between oxygen saturation values measured at different times (secondary endpoints 7 & 8), it may be appropriate to plot saturation values over time for visualisation purposes.

The two endpoints defined as time periods are not suitable for analysis with Bland-Altman plots. For each oximeter, these will be summarised using the median and quartiles. In addition, linear regression will be used to assess any correlation of these delays with the oxygen saturation value from the reference oximeter measured at the 120 second time point.

##### 11.5. The Number of Participants

The following sample size calculation was developed following discussion with experienced statisticians in the Department of Primary Health Care, and approved by a senior statistician (Dr Rafael Perera).
To calculate a sample size for a study of this type, it is necessary to define the desired accuracy of the limits of agreement (these are the ± 2SD lines on the Bland-Altman plots). As these are calculated from the sample, a larger sample will result in a smaller standard error for the limits of agreement, and hence greater confidence (smaller confidence interval) in their value. The method used in this calculation is that described in [4].
The standard deviation of the agreement between $SaO_2$ and $SpO_2$ (s) is estimated at 2.5%. This is based on estimates of 2.1% and 3.1% from the literature [5,6] and the usual quoted accuracy of ±1-2%.
Assuming an accuracy of ±1% is required for the 95% confidence intervals of the limits of agreement (i.e. the ±2SD lines on the Bland-Altman plot will have 95% confidence intervals which extend 1% in either direction), a minimum sample size of 75 participants will be required.
In addition to the minimum sample size, it is necessary to ensure an appropriate spread across the clinically relevant range of oxygen saturation values. Therefore, we intend to recruit between 20 and 25 participants with oxygen saturations in the range 96-100. To be included in this group, a participant must have reference oximeter readings within this range at the 120 second measurement time point, for both the first and second oximetry measurements.

##### 11.6. Procedure for Accounting for Missing, Unused, and Spurious Data.

Missing data will not be imputed.

Suspected spurious or anomalous data will not be removed from the data set. Bland-Altman plots allow for identification of spurious or anomalous data points, and sensitivity analysis will be carried out if such data are identified.

##### 11.7. Inclusion in Analysis

All eligible consented participants (including those with partial data) will be included in the analysis, unless they have withdrawn from the study.

##### 11.8. Procedures for Reporting any Deviation(s) from the Original Statistical Plan

Any deviations from the original statistical plan as documented in this protocol must be reported and justified in any resulting output.

© Copyright: The University of Oxford and Oxford University Hospitals NHS Trust 2014

### 12. DATA MANAGEMENT

#### 12.1. Source Data

The following data will be recorded for each participant.

On the case report form (CRF):

- Study number
- Whether consent to video recording publication has been obtained
- Date and time of start of video recording
- Randomisation protocol
- Radial pulse measured during first measurement period
- Oxygen saturation at 120s on reference oximeter during first measurement period
- Blood gas results (if measured) – SaO2
- Radial pulse measured during second measurement period
- Oxygen saturation at 120s on reference oximeter during second measurement period
- Supplemental oxygen (if present)
    - litres per minute
    - delivery method
    - estimated FiO2

On the video reporting form (duplicated by two observers):

- Study number
- Date and time of start of video recording
- Indication of skin colour (light, intermediate, dark)
- For each oximeter device and each measurement period:
    - Time at which the device is placed on the finger (defined as when the researcher is no longer touching it)
    - Time at which the device first displays oxygen saturation reading (t1)
    - Oxygen saturation reading displayed at time t1
    - Time at which the device first displays heart rate reading (t2)
    - Heart rate reading displayed at time t2
    - Time at which the device first displays “good quality” reading (judged according to manufacturers’ instructions e.g. by observing a pulse strength indicator) (t3)
    - Oxygen saturation reading displayed at time t3
    - Heart rate readings displayed at time t3
    - Oxygen saturation reading displayed 15s from the time the last device is placed
    - Oxygen saturation reading displayed 30s from the time the last device is placed
    - Oxygen saturation reading displayed 45s from the time the last device is placed
    - Oxygen saturation reading displayed 60s from the time the last device is placed
    - Oxygen saturation reading displayed 120s from the time the last device is placed
    - Heart rate reading displayed 15s from the time the last device is placed
    - Heart rate reading displayed 30s from the time the last device is placed
    - Heart rate reading displayed 45s from the time the last device is placed
    - Heart rate reading displayed 60s from the time the last device is placed

© Copyright: The University of Oxford and Oxford University Hospitals NHS Trust 2014

- Heart rate reading displayed 120s from the time the last device is placed

A study consent form, and (optionally) a consent for video recording publication form will also be obtained for each participant. These will contain the study number, and the participants name and signature, as well as the name and signature of the witnessing researcher. These will not be considered source data, and will not be used in further analysis.

##### 12.2. Access to Data

Direct access will be granted to authorised representatives from the Sponsor, host institution and the regulatory authorities to permit study-related monitoring, audits and inspections.

##### 12.3. Data Recording and Record Keeping

No personally identifiable data will be collected as part of this study, with the exception of names and signatures of participants on consent forms.

All source data will be anonymised using the study number at the time of collection, and will remain anonymised throughout the study. No list relating study numbers to participants will be held.

The participants will be identified by a unique study specific number and/or code in any database. The name and any other identifying detail will NOT be included in any study data electronic file.

**Data storage**

Paper forms will be stored in locked cabinets within offices that are only accessible to staff members employed by either the University of Oxford Department of Primary Care Health Sciences, or the Oxford University Hospitals NHS Trust. If necessary, paper forms may be archived to secure archival facilities.

Data from case report forms and video reporting forms will be transcribed into electronic form. Video recordings for which consent for publication has been obtained will also be stored in electronic form. All of the above will be identified by study number only. Data will be stored on University of Oxford and/or Oxford University Hospital NHS Trust computers, and access to data will be limited by appropriate user controls including password protection. Laptops on which data is stored will additionally be provided with full-disk encryption.

Access to original source documents (consent forms, case report forms and video reporting forms), and to video recordings of participants who have not given consent for publication will be limited to members of the research team with a legitimate need to consult the documents, and to authorised representatives for the purposes of study-related monitoring, ethical review and regulatory inspections. Source documents will be securely stored for 5 years following the completion of the study, and will then be securely destroyed.

#### 13. QUALITY ASSURANCE PROCEDURES

The study may be monitored, or audited in accordance with the current approved protocol, ICH GCP, relevant regulations and standard operating procedures.

© Copyright: The University of Oxford and Oxford University Hospitals NHS Trust 2014

#### 14. ETHICAL AND REGULATORY CONSIDERATIONS

##### 14.1. Declaration of Helsinki

The Investigator will ensure that this study is conducted in accordance with the principles of the Declaration of Helsinki.

##### 14.2. ICH Guidelines for Good Clinical Practice

The Investigator will ensure that this study is conducted in full conformity with relevant regulations and with the ICH Guidelines for Good Clinical Practice (CPMP/ICH/135/95) July 1996.

##### 14.3. Approvals

The protocol, informed consent form, participant information sheet and any proposed advertising material will be submitted to an appropriate Research Ethics Committee (REC), regulatory authorities (MHRA in the UK), and host institution(s) for written approval.

The Investigator will submit and, where necessary, obtain approval from the above parties for all substantial amendments to the original approved documents.

##### 14.4. Reporting

The CI shall submit once a year throughout the study, or on request, an Annual Progress Report to the REC, host organisation and Sponsor. In addition, an End of Study notification and final report will be submitted to the REC, host organisation and Sponsor.

##### 14.5. Participant Confidentiality

The study staff will ensure that the participants' anonymity is maintained. The participants will be identified only by a participants ID number on the CRF and any electronic database. All documents will be stored securely and only accessible by study staff and authorised personnel. The study will comply with the Data Protection Act, which requires data to be anonymised as soon as it is practical to do so.

##### 14.6. Other Ethical Considerations

This study involves attaching multiple pulse oximeters to a participant, recording the readings using a video camera, and comparing the results to that of a blood gas measurement, where this is available.

Blood samples for blood gas measurement will only be taken where this is clinically indicated as part of the participant's usual care. Written informed consent for the use of the results of these for research purposes will be obtained as part of the recruitment process.

The attachment of a single pulse oximeter is part of normal care for patients interacting with many areas of the health service, and does not pose a risk to the participant. There are no anticipated additional risks associated with the use of multiple pulse oximeters.

The main ethical issue associated with this study is the use of video recording, which could compromise a participant's anonymity. To mitigate this, the video recording device will be set up to ensure that no

© Copyright: The University of Oxford and Oxford University Hospitals NHS Trust 2014

identifiable details such as the face are visible in the video recording. Written informed consent for video recording of the participants' hands will be obtained as part of the recruitment process. To assist with publication of the results, separate written informed consent will also be sought for publication of anonymised video clips or stills. However, consent to publication will not be required for inclusion in the study.

Ethical approval will be sought from an NHS REC prior to starting recruitment.

All members of the research team will be trained in Good Clinical Practice prior to the start of recruitment, and all staff involved in data collection will either hold an NHS contract (including honorary contracts) or a valid Research Passport.

#### 15. FINANCE AND INSURANCE

##### 15.1. Funding

This study is funded by a Scientific Foundation Board grant (SFB 2013-27) from the Royal College of General Practitioners.

##### 15.2. Insurance

The University has a specialist insurance policy in place which would operate in the event of any participant suffering harm as a result of their involvement in the research (Newline Underwriting Management Ltd, at Lloyd’s of London). NHS indemnity operates in respect of the clinical treatment which is provided.

#### 16. PUBLICATION POLICY

Outputs of the research will be published in peer-reviewed journals, and may be presented at peer-reviewed conferences. All outputs should be circulated to the investigators prior to submission.

##### Authorship

All people who fulfil ICJME criteria for authorship of an output should be offered authorship:

“Authorship credit should be based on 1) substantial contributions to conception and design, acquisition of data, or analysis and interpretation of data; 2) drafting the article or revising it critically for important intellectual content; and 3) final approval of the version to be published. Authors should meet conditions 1, 2, and 3.”

It would normally be expected that the investigators listed in this document would fulfil these criteria for most or all outputs of this study. Authorship order should be agreed early in the process of drafting an output.

##### Acknowledgements

Investigators who do not qualify for authorship of an output must be acknowledged. Appropriate acknowledgement of funding must also be ensured.

Researchers who fulfil one or more ICJME authorship criterion, but do not fulfil all three criteria should be acknowledged in journal papers and oral presentations.

© Copyright: The University of Oxford and Oxford University Hospitals NHS Trust 2014

##### Informing participants

Direct informing of participants is precluded due to anonymisation. However, steps should be taken to publicise the results of the study in such a way that participants may find out about it (e.g. via leaflets or posters in outpatients COPD clinics, and via email to the same lists from which healthy participants were recruited).

© Copyright: The University of Oxford and Oxford University Hospitals NHS Trust 2014

#### 18. APPENDIX A: AMENDMENT HISTORY

| Amendment No. | Protocol Version No. | Date issued | Author(s) of changes | Details of Changes made |
|---|---|---|---|---|
| 1 | 1.6 | 12 Jan 2014 | S Fleming | Change of end date from March to May |

Protocol amendments must be submitted to the Sponsor for approval prior to submission to the REC committee or MHRA.

#### 19. Appendix B: Equation for 95% CI of sample standard deviation

This uses the chi-squared distribution with n-1 degrees of freedom.

$$\sqrt{\frac{(n-1)s^2}{\chi^2_{0.025}}} \leq \sigma \leq \sqrt{\frac{(n-1)s^2}{\chi^2_{0.975}}}$$

© Copyright: The University of Oxford and Oxford University Hospitals NHS Trust 2014
