## Supplemental for "Accuracy of fingertip pulse oximeters: a device accuracy study"

**Supplementary data**

Supplementary Figure S1: The test pulse oximeters

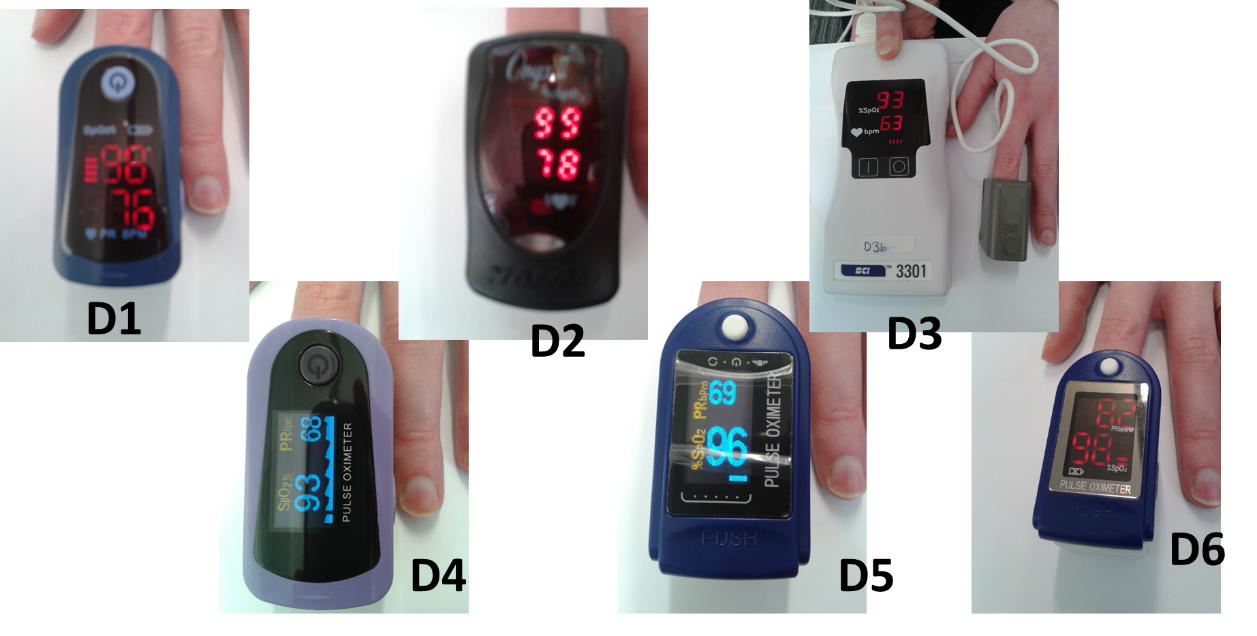

Supplementary Table S1: Identification and purchase price of test pulse oximeters

| Oximeter | Make/Model | Cost (2014) |
| --- | --- | --- |
| D1 | Merlin M-pulse lite | £49 |
| D2 | Nonin Onyx II 9550 | £229 |
| D3 | BCI 3301 | £284.96 |
| D4 | Choicemed MD300C23 | £69.50 |
| D5 | Contec CMS50D | £37.50 |
| D6 | Contec CMS50DL | £22.95 |

Supplementary Table S2: Inclusion and exclusion criteria

| **Inclusion Criteria** | **Exclusion Criteria** |
| --- | --- |
| Willing and able to give informed consent | Current application of dyes, false nails, nail polish, or other cosmetics (e.g. henna) on the fingers of the right hand. |
| Aged 18 years or above | Amputation of any part of the right upper limb resulting in loss of one or more fingers on the right hand. |
| Fulfils one of the following two criteria:   - current patient due to attend the respiratory outpatients clinic during the study period - adult who is in good general health. | Recent (<1 week) medical procedure involving the administration of intravenous dyes or contrast media. |

Supplementary Table S3: Number of data points from the double-extracted video data that met the criteria for inclusion in data analysis, either due to complete agreement, or close agreement. The total number of data points for each measurement was 600 (75 patients x 4 oximeters x 2 measurement sessions.) Overall measurements were assessed excluding the first good quality value, as this was more subjective.

| **Data** | **Number (%) meeting inclusion criteria** | **Number (%) in complete agreement** | **Number (%) averaged due to close agreement** |
| --- | --- | --- | --- |
| **Oxygen Saturation (SpO_2_)** | | | |
| First value | 524 (87.3%) | 518 (86.3%) | 6 (1%) |
| First good quality value* | 511 (85.2%) | 501 (83.5%) | 10 (1.7%) |
| 15 seconds | 517 (86.2%) | 478 (79.7%) | 39 (6.5%) |
| 30 seconds | 543 (90.5%) | 499 (83.2%) | 44 (7.3%) |
| 45 seconds | 552 (92%) | 512 (85.3%) | 40 (6.7%) |
| 60 seconds | 553 (92.2%) | 513 (85.5%) | 40 (6.7%) |
| 120 seconds | 552 (92%) | 499 (83.2%) | 53 (8.8%) |
| Overall (N=3600) | 3241 (90%) | 3019 (83.9%) | 222 (6.2%) |
| **Heart rate** | | | |
| First value | 500 (83.3%) | 456 (76%) | 44 (7.3%) |
| First good quality value | 484 (80.7%) | 386 (64.3%) | 98 (16.3%) |
| 15 seconds | 520 (86.7%) | 368 (61.3%) | 152 (25.3%) |
| 30 seconds | 542 (90.3%) | 415 (69.2%) | 127 (21.2%) |
| 45 seconds | 544 (90.7%) | 409 (68.2%) | 135 (22.5%) |
| 60 seconds | 542 (90.3%) | 376 (62.7%) | 166 (27.7%) |
| 120 seconds | 543 (90.5%) | 377 (62.8%) | 166 (27.7%) |
| Overall (N=3600) | 3191 (88.6%) | 2401 (66.7%) | 790 (21.9%) |
| **Times** | | | |
| Time attached | 588 (98%) | 324 (54%) | 264 (44%) |
| Time of first good quality signal | 408 (68%) | 68 (11.3%) | 340 (56.7%) |

* The first good quality value was greater than 15 seconds in 15/600 measurements

upplementary Table S4: Results of Bland-Altman analysis (Bias and 95% limits of agreement) plus 95% confidence intervals for each oximetry device measured at 2 minutes. The reference oximeter is denoted as “R”. In each case differences were calculated as the references (SaO_2_, or reference SpO_2_) minus the value displayed on the test oximeter.

| Device | SpO2 compared to reference device | | SpO2 compared to blood gas analysis | |
| --- | --- | --- | --- | --- |
|  | Bias (95% CI) | 95% LOA (95% CI) | Bias (95% CI) | 95% LOA (95% CI) |
| D1 | -2.4% (-1.8 to -3.0) | -7.4% (-8.5 to -6.3) to 2.6% (1.5 to 3.7) | -0.5% (-1.4 to 0.4) | -5.8% (-7.3 to -4.3) to 4.8% (3.3 to 6.3) |
| D2 | -1.6% (-0.9 to -2.3) | -7.1% (-8.3 to -5.9) to 3.9% (2.7 to 5.1) | 0.7% (-0.2 to 1.7) | -4.9% (-6.5 to -3.3) to 6.4% (4.8 to 8.0) |
| D3 | -0.6% (-0.1 to -1.2) | -4.9% (-5.8 to -4.0) to 3.6% (2.7 to 4.6) | 1.4% (0.6 to 2.1) | -3.5% (-4.8 to -2.2) to 6.2% (4.9 to 7.5) |
| D4 | -2.6% (-2.1 to -3.1) | -6.5% (-7.3 to -5.6) to 1.3% (0.5 to 2.1) | -1.2% (-1.8 to -0.6) | -5.0% (-6.0 to -3.9) to 2.6% (1.6 to 3.7) |
| D5 | -1.1% (-0.5 to -1.7) | -5.8% (-6.9 to -4.8) to 3.6% (2.6 to 4.7) | 0.8% (-0.2 to 1.7) | -4.8% (-6.4 to -3.2) to 6.3% (4.7 to 7.9) |
| D6 | -2.4% (-1.7 to -3.0) | -7.4% (-8.4 to -6.3) to 2.7% (1.6 to 3.7) | -0.6% (-1.4 to 0.3) | -5.9% (-7.4 to -4.4) to 4.8% (3.3 to 6.3) |
| R | N/A | N/A | 2.3 % (1.5 to 3.0) | -4.1% (-5.3 to -2.8) to 8.6% (7.3 to 9.8) |

LOA: Limits of agreement

Supplementary Table S5: Results of Bland-Altman analysis (Bias and 95% limits of agreement) plus 95% confidence intervals for heart rate measured by each oximetry device measured at 2 minutes and 30 seconds. The reference oximeter is denoted as “R”. In each case differences were calculated as the reference (manual heart rate) minus the value displayed on the test oximeter.

| Device | Heart rate at 2 minutes compared to manual pulse | | Heart rate at 30 seconds compared to manual pulse | |
| --- | --- | --- | --- | --- |
|  | Bias (95% CI) | 95% LOA (95% CI) | Bias (95% CI) | 95% LOA (95% CI) |
| D1 | -1.9bpm (-3.2 to -0.6) | -12.6bpm (-14.8 to -10.4) to 8.7bpm (6.5 to 10.9) | -0.3bpm (-1.9 to 1.3) | -13.1bpm (-15.9 to -10.4) to 12.5bpm (9.7 to 15.2) |
| D2 | -2.2bpm (-3.8 to -0.5) | -15.6bpm (-18.5 to -12.8) to 11.3bpm (8.5 to 14.1) | -2.1bpm (-3.9 to -0.2) | -17.4bpm (-20.6 to -14.2) to 13.2bpm (10.1 to 16.5) |
| D3 | -0.9bpm (-2.4 to 0.5) | -12.9bpm (-15.3 to -10.4) to 11.0bpm (8.5 to 13.5) | -0.1bpm (-2.6 to 2.3) | -20.3bpm (-24.5 to -16.2) to 20.1bpm (15.9 to 24.3) |
| D4 | -1.9bpm (-3.4 to -0.4) | -13.9bpm (-16.5 to -11.3) to 10.1bpm (7.5 to 12.7) | -1.9bpm (-3.1 to -0.7) | -11.4bpm (-13.5 to -9.4) to 7.6bpm (5.5 to 9.7) |
| D5 | -0.8bpm (-3.0 to 1.4) | -16.9bpm (-20.7 to -13.2) to 15.4bpm (11.7 to 19.2) | -1.3bpm (-3.4 to 0.8) | -16.8bpm (-20.5 to -13.2) to 14.2bpm (10.5 to 17.8) |
| D6 | -2.7bpm (-4.1 to -1.2) | -14.8bpm (-17.3 to -12.3) to 9.5bpm 6.9 to 12.0) | -1.8bpm (-2.9 to -0.6) | -11.4bpm (-13.3 to -9.4) to 7.8bpm (5.8 to 9.8) |
| R | -1.6bpm (-2.6 to -0.7) | -12.5bpm (-14.1 to -11.0) to 9.3bpm (7.7 to 10.9) | -1.6bpm (-3.0 to -0.2) | -18.7bpm (-21.2 to -16.3) to 15.5bpm (13.1 to 8.0) |

LOA: Limits of agreement

Supplementary Table S6: Results of Bland-Altman analysis (Bias and 95% limits of agreement) for each oximetry device measured at 2 minutes after exclusion of data from one patient with very low oxygen saturation. The reference oximeter is denoted as “R”. In each case differences were calculated as the references (SaO_2_, reference SpO_2_) minus the value displayed on the test oximeter.

| Device | Cost | Bias (95% LOA) for SpO_2_ compared to reference device | Bias (95% LOA) for SpO_2_ compared to blood gas analysis |
| --- | --- | --- | --- |
| D1 | £49 | -2.2% (-5.6 to 1.3) | -0.4 %(-5.7 to 4.8) |
| D2 | £229 | -1.3% (-3.6 to 1.0) | 0.9% (-4.1 to 6.0) |
| D3 | £284.96 | -0.4% (-2.8 to 2.0) | 1.4% (-3.4 to 6.2) |
| D4 | £69.50 | -2.4% (-4.9 to 0.0) | -1.2% (-5.0 to 2.6) |
| D5 | £37.50 | -0.8% (-3.7 to 2.0) | 0.8% (-4.7 to 6.4) |
| D6 | £22.95 | -2.1% (-5.3 to 1.0) | -0.5% (-5.8 to 4.8) |
| R |  | N/A | 1.9% (-3.2 to 7.1) |

LOA: Limits of agreement

Supplementary Table S7: Results of Bland-Altman analysis (bias and 95% limits of agreement) for errors of SpO_2_ compared to the value at 120 seconds for each oximetry device at 6 different time points. In each case differences were calculated as the SpO_2_ at 120 seconds minus the value at the time point of interest.

| Time | D1 | D2 | D3 | D4 | D5 | D6 | R |
| --- | --- | --- | --- | --- | --- | --- | --- |
| First SpO_2_ | -0.37%  (-3.61 to 2.86) | -0.43%  (-3.23 to 2.36) | -0.74%  (-3.3 to 1.82) | -0.83%  (-4.47 to 2.81) | 0.50%  (-2.63 to 3.63) | 0.84%  (-2.91 to 4.60) | 0.16%  (-10.1 to 10.4) |
| First good quality SpO_2_ | -0.34%  (-3.24 to 2.56) | -0.46%  (-3.29 to 2.37) | -0.75%  (-3.32 to 1.82) | -0.82%  (-4.42 to 2.78) | 0.44%  (-2.69 to 3.57) | 0.86%  (-2.92 to 4.64) | 0.15%  (-10.2 to 10.5) |
| 15 seconds | -0.29%  (-2.79 to 2.21) | -0.14%  (-1.94 to 1.66) | -0.33%  (-2.47 to 1.80) | -0.57% (-3.60 to 2.45) | -0.052%  (-2.27 to 2.17) | 0.19%  (-2.89 to 3.27) | -0.12%  (-3.16 to 2.77) |
| 30 seconds | -0.094%  (-1.80 to 1.62) | -0.14%  (-2.02 to 1.75) | -0.29%  (-2.48 to 1.91) | -0.50%  (-2.82 to 1.82) | -0.086%  (-1.82 to 1.65) | -0.046%  (-2.84 to 2.75) | -0.23%  (-2.25 to 1.79) |
| 45 seconds | -0.052%  (-1.68 to 1.57) | -0.086%  (-1.89 to 1.72) | -0.37%  (-2.91 to 2.17) | -0.43%  (-2.71 to 1.86) | -0.11%  (-1.86 to 1.65) | -0.27%  (-2.61 to 2.07) | -0.16%  (-2.24 to 1.92) |
| 60 seconds | -0.022%  (-1.66 to 1.62) | -0.034%  (-1.55 to 1.49) | -0.29%  (-2.75 to 2.18) | -0.33%  (-2.27 to 1.62) | -0.22%  (-1.94 to 1.51) | -0.13%  (-2.48 to 2.22) | -0.27%  (-2.19 to 1.64) |

Supplementary Table S8: Results of Bland-Altman analysis (Bias and 95% limits of agreement) for each oximetry device after 30 seconds. In each case differences were calculated as the reference oximeter minus the value displayed on the test oximeter.

| Device | Bias (95% LOA) for SaO_2_ compared to blood gas analysis | |
| --- | --- | --- |
|  | Bias (95% CI) | 95% LOA (95% CI) |
| D1 | -2.6% (-3.3 to -2.0) | -7.6% (-8.8 to -6.5) to 2.4% (1.3 to 3.5) |
| D2 | -1.3% (-2.1 to -0.6) | -7.5 (-8.9 to -6.2) to 4.8% (3.5 to 6.1) |
| D3 | -0.3% (-0.8 to 0.2) | -4.4 (-5.3 to -3.5) to 3.7% (2.9 to 4.6) |
| D4 | -2.0% (-2.3 to -1.6) | -4.5 (-5.1 to -4.0) to 0.6% (0.0 to 1.2) |
| D5 | -0.9% (-1.6 to -0.2) | -6.2 (-7.4 to -5.0) to 4.4% (3.2 to 5.6) |
| D6 | -2.3% (-3.0 to -1.6) | -8.2 (-9.5 to -6.9) to 3.6% (2.3 to 4.9) |

LOA: Limits of agreement

Supplementary Table S9: Results of linear regression for time to first good quality value (in seconds), by oxygen saturation at 2 minutes, including adjustment for device type.

|  | Univariate Model | | | Adjusted for device type | | |
| --- | --- | --- | --- | --- | --- | --- |
|  | Estimate | 95% CI | P-value | Estimate | 95% CI | P-value |
| SpO_2_ at 2 minutes (%) | 0.014 | -0.051 to 0.078 | 0.678 | 0.030 | -0.023 to 0.083 | 0.260 |
| D1 (Reference) |  |  |  | 1 |  |  |
| D2 |  |  |  | -0.405 | -2.036 to 1.226 | 0.625 |
| D3 |  |  |  | 3.312 | 1.710 to 4.914 | <0.001 |
| D4 |  |  |  | -0.905 | -2.517 to 0.707 | 0.270 |
| D5 |  |  |  | 3.635 | 1.926 to 5.344 | <0.001 |
| D6 |  |  |  | 2.798 | 1.112 to 4.483 | 0.001 |
| R |  |  |  | 8.105 | 6.575 to 9.634 | <0.001 |
